# Sex-specific prediction of major cardiovascular events in apparently healthy individuals with multi-omics data

**DOI:** 10.64898/2026.02.19.26346632

**Authors:** Ruijie Xie, Megha Bhardwaj, Sha Sha, Lei Peng, Tomislav Vlaski, Hermann Brenner, Ben Schöttker

## Abstract

**Background:** While multi-omics approaches, incorporating polygenic risk scores (PRS), metabolomics, and proteomics have shown promise in predicting major adverse cardiovascular events (MACE), their added value beyond cardiovascular disease (CVD) risk factors remains underexplored. We aimed to assess whether integrating multi-omics biomarkers into the SCORE2 model improves the prediction of MACE in apparently healthy individuals.

**Methods:** This study included 24,042 UK Biobank participants without CVD or diabetes mellitus, aged 40-69 years. Multi-omics biomarkers were fitted in sex-specific models including the variables of SCORE2 and 9 metabolites, 12 proteins, and a PRS for CVD in males, as well as 7 metabolites, 11 proteins, and a PRS for CVD in females. The performance of the SCORE2 model and its multi-omics extensions was compared using Harrell’s C-index and the net reclassification index (NRI) in a training and test set (70% and 30% of study population).

**Results:** In 10-year follow-up, 1,204 MACE events occurred. Integrating multi-omics biomarkers into SCORE2 significantly improved the predictive performance (C-index: 0.708 to 0.769, *P*<0.001; NRI=26.2%). In males, the C-index improved from 0.682 to 0.752 (ΔC-index=+0.070, *P*<0.001; NRI=12.4%), while in females, it increased from 0.724 to 0.782 (ΔC-index=+0.058, *P*<0.001; NRI=30.4%). However, full multi-omics measurements may not be needed because the combination of proteomics and PRS yielded comparable performance in males (C-index=0.749) and females (C-index=0.782).

**Conclusions:** Integrating a protein panel and a PRS significantly improves MACE risk prediction by the SCORE2 model, which includes HDL and total cholesterol. Adding further metabolites has limited additional predictive value.

## Introduction

Cardiovascular disease (CVD) remains a leading cause of morbidity and mortality worldwide, presenting a significant global health challenge [1]. Despite advancements in risk prediction and management, such as the development of SCORE2 in 2021 to estimate 10-year major adverse cardiovascular events (MACE) risk for individuals without CVD or diabetes, there remains a critical need to improve precision in risk assessment [2, 3].

Omics-based research, encompassing genomics, proteomics, and metabolomics, enhances CVD risk prediction by offering a more comprehensive measurement of disease-related biological processes than captured in traditional CVD risk factor models [4]. Genome-wide association studies (GWAS) have identified numerous genetic risk variants for CVD, while proteomics and metabolomics have illuminated key pathways related to inflammation, lipid metabolism, and vascular function [5–7]. Early omics studies have primarily focused on the predictive value of single omics layers in isolation, demonstrating modest but statistically significant improvements in CVD risk prediction [8–10]. Nonetheless, the extent to which combining these layers enhances predictive value over individual omics layers remains insufficiently studied [11], primarily due to the high costs for these measurements. The recent establishment of multi-omics measurements within a large sub-sample of the UK Biobank (UKB) cohort now provides a unique resource to systematically investigate the added value of integrative omics approaches at population scale [12].

In previous research with the UKB from our group, a proteomics based as well as a metabolomics based MACE risk score were developed that significantly enhance the predictive value of the SCORE2 model [13, 14]. SCORE2 is a traditional sex-specific risk factor model used by the European Society of Cardiology to assess the 10-year MACE risk based on age, smoking, systolic blood pressure (SBP), high-density lipoprotein cholesterol (HDL-C) and total cholesterol (TC). In addition, a polygenic risk score (PRS) for CVD prediction has also been developed in the UKB [15].

We aim to build upon this previous research on the single omics layers and combine the developed risk algorithms for proteomics [13], metabolomics [14], and GWAS data [15] with the SCORE2 model [2] and assess which combination of omics data leads to the optimal multi-omics based 10-year MACE prediction model. As emerging evidence underscores significant sex differences in CVD pathophysiology [16, 17], we strive for sex-specific prediction models.

## Methods

### Study population

The UKB is a large prospective cohort with 502,414 participants, aged 37 to 73 years, recruited from 13 March 2006 to 1 October 2010 across 22 assessment sites in England, Scotland, and Wales [18]. The cohort features an extensive collection of deep phenotypic and genetic data, including blood and urine biomarkers, whole-body imaging, lifestyle factors, physical and anthropometric measurements, genome-wide genotyping, and exome and genome sequencing.

The starting point for the sample selection was those n=54,219 UKB participants with available proteomics measurements (see study flow-chart in **Figure 1**). We excluded participants with more than 50% missing protein measurements (n=1,871), and subjects who were not randomly selected and did not develop MACE during follow-up (n=5,282). Due to the resulting oversampling of n=1,204 MACE cases, which were not part of the random sample, the study design is a case-cohort design [19]. This study design has high statistical power while maintaining the cohort design excluding any risk of selection bias.

**Figure 1.**
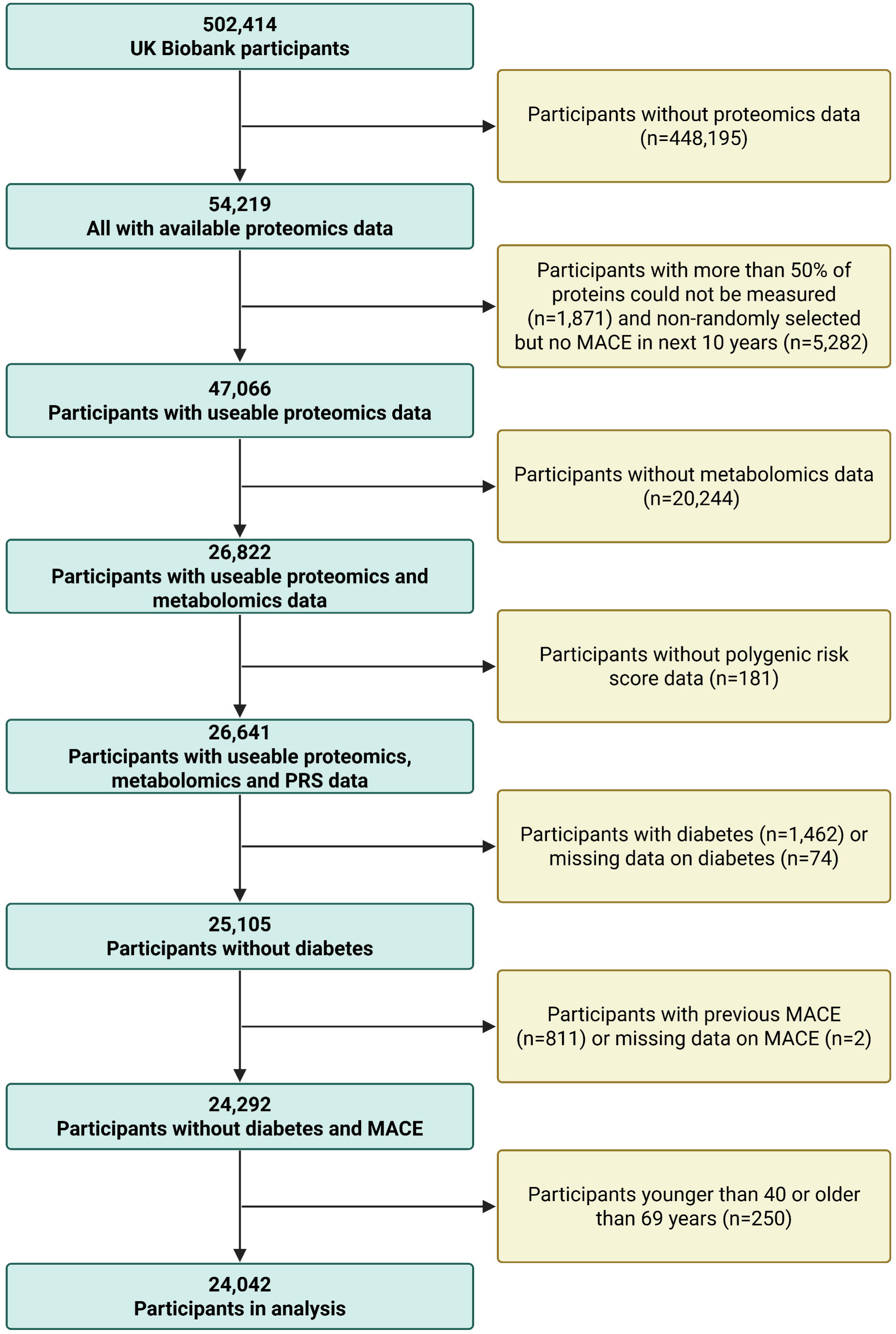
Flowchart of participant selection **Abbreviations**: MACE, major adverse cardiovascular events.

Subsequently, participants lacking metabolomics data (n=20,244), or polygenic risk score (PRS) data (n=181) were excluded, resulting in n=26,641 participants with complete multi-omics data. Furthermore, we excluded participants with diabetes (n=1,462) or missing diabetes status (n=74), those with prior MACE (n=811) or missing MACE data (n=2), and participants younger than 40 or older than 69 years (n=250). Ultimately, the final analysis comprised 24,042 participants.

### Variables of the SCORE2 model

The SCORE2 model, designed for adults without diabetes aged 40 to 69, includes age, sex, smoking status, SBP, HDL-C, and TC [2]. Data on age, sex, and smoking status were obtained through standardized questionnaires. HDL-C and total cholesterol levels were measured using the Beckman Coulter AU5800 with an enzymatic method. SBP was recorded through automated readings using the Omron device on the left upper arm.

### Proteomics data

Proteomic profiling was conducted on EDTA-plasma samples collected at baseline. The assay protocols, including sample handling and selection procedures, have been described previously [20]. In brief, a total of 2,923 unique proteins were measured using the Olink Explore 3072 platform (Olink Proteomics, Uppsala, Sweden). Olink utilizes the Proximity Extension Assay (PEA) method that targets proteins via pairs of antibodies linked to complementary oligonucleotides [21–23]. As outlined in our prior study [24], proteins with more than 20% missing values or with over 25% of values below the detection limit were excluded (n = 838). Ultimately, 2,085 proteins from the Olink Explore platform were included in the proteomic biomarker selection process. In our previous study [13], we applied sex-specific least absolute shrinkage and selection operator (LASSO) regression to identify a parsimonious signature of 18 proteins (5 common to both sexes, 7 male-specific and 6 female-specific; **Supplemental Table S1**). Incorporation of these proteins into the SCORE2 model substantially improved discrimination: The C-index increased from 0.713 to 0.778 in the total population (*P*<0.001), with ΔC-index=+0.087 in men (from 0.684 to 0.771) and ΔC-index=+0.049 in women (from 0.720 to 0.769). In the present analysis, we evaluated only these 18 pre-selected proteins.

### Metabolomic data

Nightingale Health utilized a high-throughput nuclear magnetic resonance (NMR) metabolomics platform to analyze plasma samples [25]. These biomarkers encompass a range of molecules, including lipids, fatty acids, amino acids, ketone bodies, and other critical low-molecular-weight compounds. A total of 249 metabolites were measured, of which 168 were quantified in absolute concentrations (including 61 composite biomarkers derived from 107 directly measured biomarkers), while 81 biomarkers were expressed as ratios. In our previous study [14], we applied sex-specific LASSO regression to the 249 metabolites and identified a parsimonious signature comprising 13 metabolites (3 common to both sexes, 6 male-specific and 4 female-specific; **Supplemental Table S1**). Incorporation of these metabolites into the SCORE2-Diabetes model increased C-index from 0.691 to 0.710 in the total population (*P*<0.001), with ΔC-index=+0.027 in men (from 0.638 to 0.665) and ΔC-index=+0.023 in women (from 0.686 to 0.709). In the present analysis, we evaluated only these 13 pre-selected metabolites.

### Polygenic risk score for CVD

We obtained the PRS for CVD from the UK Biobank PRS Release [15]. This pre-computed score incorporates 1.42 million genetic variants with non-zero weights. The score was developed using a Bayesian modeling framework, initially trained on meta-analyzed summary statistics from nine external GWAS datasets. These summary statistics were sourced from large-scale international consortia, primarily CARDIoGRAMplusC4D and MEGASTROKE, where genotyping was conducted on a variety of Illumina and Affymetrix arrays, with Illumina platforms being predominant [26]. Subsequently, it was optimized using individual-level data from the UK Biobank cohort, for which genotyping was conducted using the UK Biobank Axiom and UK BiLEVE Axiom arrays [15]. According to internal validation analyses, this CVD-PRS demonstrates robust predictive performance and is superior to previously published scores [15, 27].

### Outcome ascertainment

The primary endpoint was MACE, defined as an endpoint of cardiovascular death, non-fatal myocardial infarctions, and non-fatal strokes, in alignment with the SCORE2 model [2]. Occurrences of non-fatal myocardial infarctions and strokes were identified using primary care records or hospital episode statistics. Dates and causes of death were determined through death registries from the National Health Service Information Centre in England and Wales, and the National Health Service Central Register in Scotland. Participants were followed from baseline until the first occurrence of a MACE event, death, or the end of the ten-year follow-up period, whichever occurred first. Non-cardiovascular deaths were treated as competing risks in the analysis. A comprehensive definition of MACE is available in **Supplemental Table S2**.

### Statistical analyses

#### General remarks

All analyses were performed using R software (version 4.4.0, R Foundation for Statistical Computing, Vienna, Austria). Statistical significance was defined as *P*-values < 0.05 for two-sided tests. Missing values of variables of the SCORE2 (variable with the highest proportion of missing values was HDL-C with 12.2%) and multi-omics (mostly complete with a few proteins with up to 20% of missing values) were single imputed using the chained equations method with random forest algorithms implemented in the R package *missForest* (version 1.5) [28].

#### Test of Model Performance

The study design is illustrated in **Figure 2**. Protein panels (12 proteins for men and 11 for women), metabolite panels (9 metabolites for men and 7 for women), and the UK Biobank’s CVD-PRS, derived in previously published selection procedures [13–15], were combined with the SCORE2 variables to create the optimal multi-omics based, sex-specific 10-year MACE risk prediction model (biomarker names listed in **Supplemental Table 1**). The UKB dataset was randomly divided into a training set (70%) and a test set (30%) separately for male and female participants. The performance of the derived models was validated using the test set (30% of the UKB).

**Figure 2.**
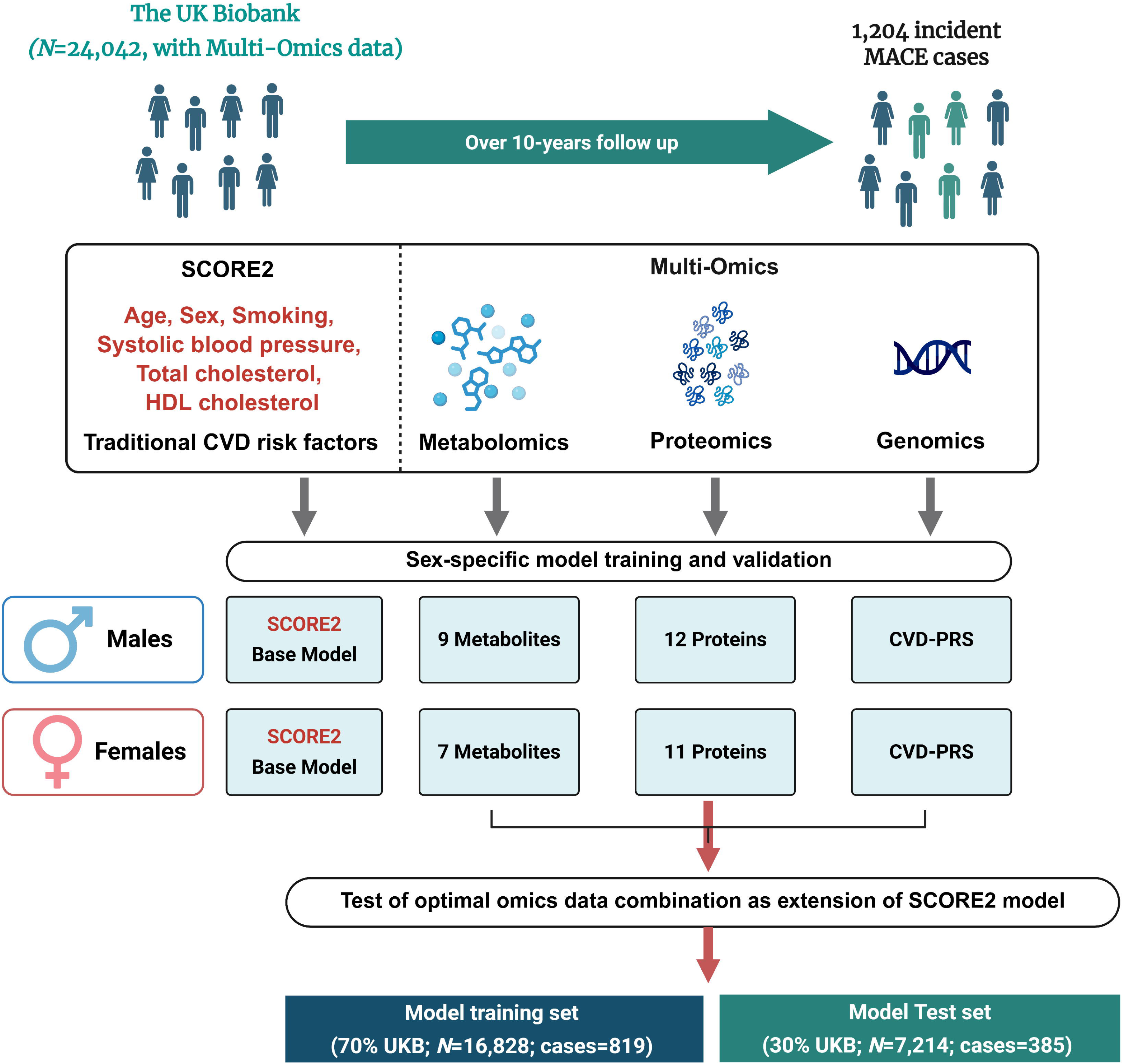
Study design **Abbreviations**: HDL, high-density lipoprotein; MACE, major adverse cardiovascular events; PRS, polygenic risk score.

In a first step, the model performance of each multi-omics layer was tested when added to the SCORE2. In a second step, a multi-omics + SCORE2 variables predictor was tested against the SCORE2 variables. In a third step, we evaluated if all multi-omics layers are needed or if some data could be removed without losing predictive power. To get a better overview, the incremental C-statistic of each single biomarker was estimated. Finally, the best performing multi-omics layers were added one by one to the SCORE 2 base model until the addition of another layer did not improve the model’s C-index any more with statistical significance. To address the competing risk of non-cardiovascular death, all models were derived using the Fine and Gray subdistribution hazard regression, implemented in the *cmprsk* R package (version 2.2-11) [29].

Model performance was primarily assessed for its discrimination using Harrell’s C-index, specifically adapted for competing risk scenarios via the the *riskRegression* R package (version 2023.03.2). The statistical significance of improvements in the C-index was evaluated using the method for comparing correlated C-indices in survival analysis proposed by Kang et al., implemented in the R package *compareC* (version 1.3.2) [30]. Risk reclassification was evaluated using the net reclassification index (NRI) and the integrated discrimination index (IDI) [31]. Pre-specified cardiovascular risk categories (0–7.5%, >7.5–15%, and >15%) were applied to calculate the NRI, assessing the proportion of individuals correctly reclassified compared to the SCORE2 model [2]. Model calibration was assessed by plotting observed MACE event rates against predicted rates across deciles of absolute predicted risk.

#### Associations of selected multi-omics biomarkers with MACE

To report hazard ratios (HRs) and 95% confidence intervals (CIs) for the associations of the tested multi-omics biomarkers (per one standard deviation increment) with 10-year MACE incidence in male and female participants, the biomarkers were individually added to Cox proportional hazards regression models in the validation set. These models were adjusted for the variables of the SCORE2 model using the *survival* package (version 3.5-5) in R.

#### Correlation matrix

In addition, Spearman correlation coefficients were calculated to evaluate the independence of the evaluated multi-omics biomarkers.

## Results

### Baseline characteristics and MACE case numbers

**Table 1** presents the baseline characteristics of the 24,042 participants included in the analysis, comprising 10,528 males (43.8%) and 13,514 females (56.2%). The overall mean age was 56.2 ± 8.1 years, with similar age distributions in males (56.1 ± 8.3 years) and females (56.3 ± 8.0 years). Compared to females, males had a higher prevalence of current smoking (12.0% vs. 8.8%), higher SBP (142.8 ± 18.4 mmHg vs. 136.7 ± 20.3 mmHg), and lower HDL-C levels (1.3 ± 0.3 mmol/L vs. 1.6 ± 0.4 mmol/L). In contrast, females had slightly higher total cholesterol levels (5.9 ± 1.1 mmol/L vs. 5.6 ± 1.0 mmol/L).

**Table 1.** Baseline characteristics of selected participants from the UK Biobank.

| **Baseline characteristics** | **Total**<br>**(N=24,042)** | **Male**<br>**(N=10,528)** | **Female**<br>**(N=13,514)** |
| --- | --- | --- | --- |
| Age (years) | 56.2 (8.1) | 56.1 (8.3) | 56.3 (8.0) |
| Current smoker, N (%) | 2,448 (10.2) | 1,262 (12.0) | 1,186 (8.8) |
| Systolic blood pressure (mmHg) | 139.4 (19.7) | 142.8 (18.4) | 136.7 (20.3) |
| Total cholesterol (mmol/L) | 5.8 (1.1) | 5.6 (1.0) | 5.9 (1.1) |
| HDL cholesterol (mmol/L) | 1.5 (0.4) | 1.3 (0.3) | 1.6 (0.4) |

Values are expressed as mean (standard deviation) for continuous variables, and as N (%) for categorical variables.
**Abbreviations:** HDL, high-density lipoprotein; MACE, major adverse cardiovascular event.

Over a 10-year follow-up period, a total of 1,204 MACE events were recorded, comprising 742 events in males and 462 in females.

### Correlations among biomarkers and associations with MACE

Figures 3A and **3B** illustrate the Spearman correlation matrices among the PRS, the selected metabolites, and proteins in males and females, respectively. Almost no correlations (all r ≤ 0.05) were observed for the CVD-PRS with metabolites or proteins in both sexes. Correlations between metabolites and proteins were also weak with the exception of GlycA (Glycoprotein acetyls), which is an inflammatory biomarker and is moderately correlated with IL6 (Interleukin-6) (r=0.33 among males) and a few other inflammation related proteins. Within metabolomic and proteomic biomarker panels, a few demonstrated strong correlations (defined as r≥0.6). In males, WFDC2 (WAP four-disulfide core domain protein) was strongly correlated with CXCL17 (C-X-C motif chemokine 17; r=0.60) and GDF15 (Growth/differentiation factor 15; r=0.61). In females, IDL-CE-pct (The cholesteryl esters percentage of total lipids in intermediate-density lipoprotein) and L-LDL-TG-pct (Triglycerides to total lipids in large low-density lipoprotein percentage) showed a strong correlation (r=0.70).

**Figure 3.**
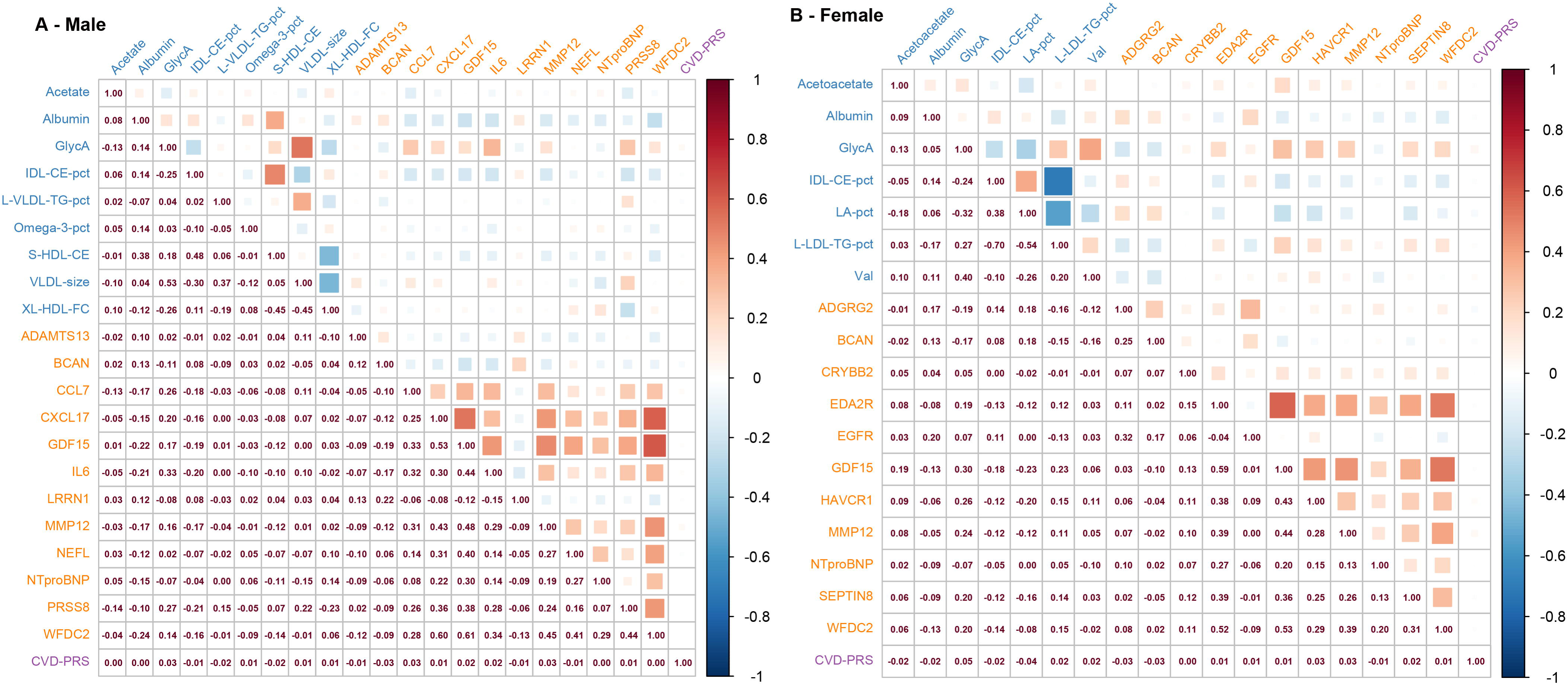
Sex-specific correlation matrices of selected biomarkers in the validation set (30% of UK Biobank, N=7,214) Spearman correlation matrices of selected PRS, metabolites, and proteins are shown for males (Panel A) and females (Panel B). **Abbreviations:** ADAMTS13, A disintegrin and metalloproteinase with thrombospondin motifs 13; BCAN, Brevican core protein; CCL7, C-C motif chemokine 7; CRYBB2, Beta-crystallin B2; CXCL17, C-X-C motif chemokine 17; EDA2R, Tumor necrosis factor receptor superfamily member 27; EGFR, Epidermal growth factor receptor; GDF15, Growth/differentiation factor 15; GlycA, Glycoprotein acetyls; IDL-CE-pct, The cholesteryl esters percentage of total lipids in intermediate-density lipoprotein; IL6, Interleukin-6; LA-pct, The linoleic acid percentage of total fatty acids; L-LDL-TG-pct, Triglycerides to total lipids in large low-density lipoprotein percentage; L-VLDL-TG-pct, The triglycerides percentage of total lipids in large very-low-density lipoprotein; LRRN1, Leucine-rich repeat neuronal protein 1; MMP12, Macrophage metalloelastase; NEFL, Neurofilament light polypeptide; NTproBNP, N-terminal prohormone of brain natriuretic peptide; Omega-3-pct, The Omega-3 fatty acids percentage of total fatty acids; PRS, polygenic risk score; PRSS8, Prostasin; S-HDL-CE, Cholesteryl esters in small high-density lipoprotein; SEPTIN8, Septin-8; Val, Valine; VLDL-size, Average diameter for very-low-density lipoprotein particles; WFDC2, WAP four-disulfide core domain protein; XL-HDL-FC, Free cholesterol in very large high-density lipoprotein.

Figures 4A and **4B** show the associations of the selected multi-omics biomarkers with MACE risk in males and females, respectively. Most biomarkers exhibited significant associations with MACE, with overlapping biomarkers in males and females demonstrating consistent effect directions. In males, five metabolites and two proteins were not significantly associated with MACE. In contrast, all biomarkers except for valine (Val) were significantly associated with MACE risk in females.

**Figure 4.**
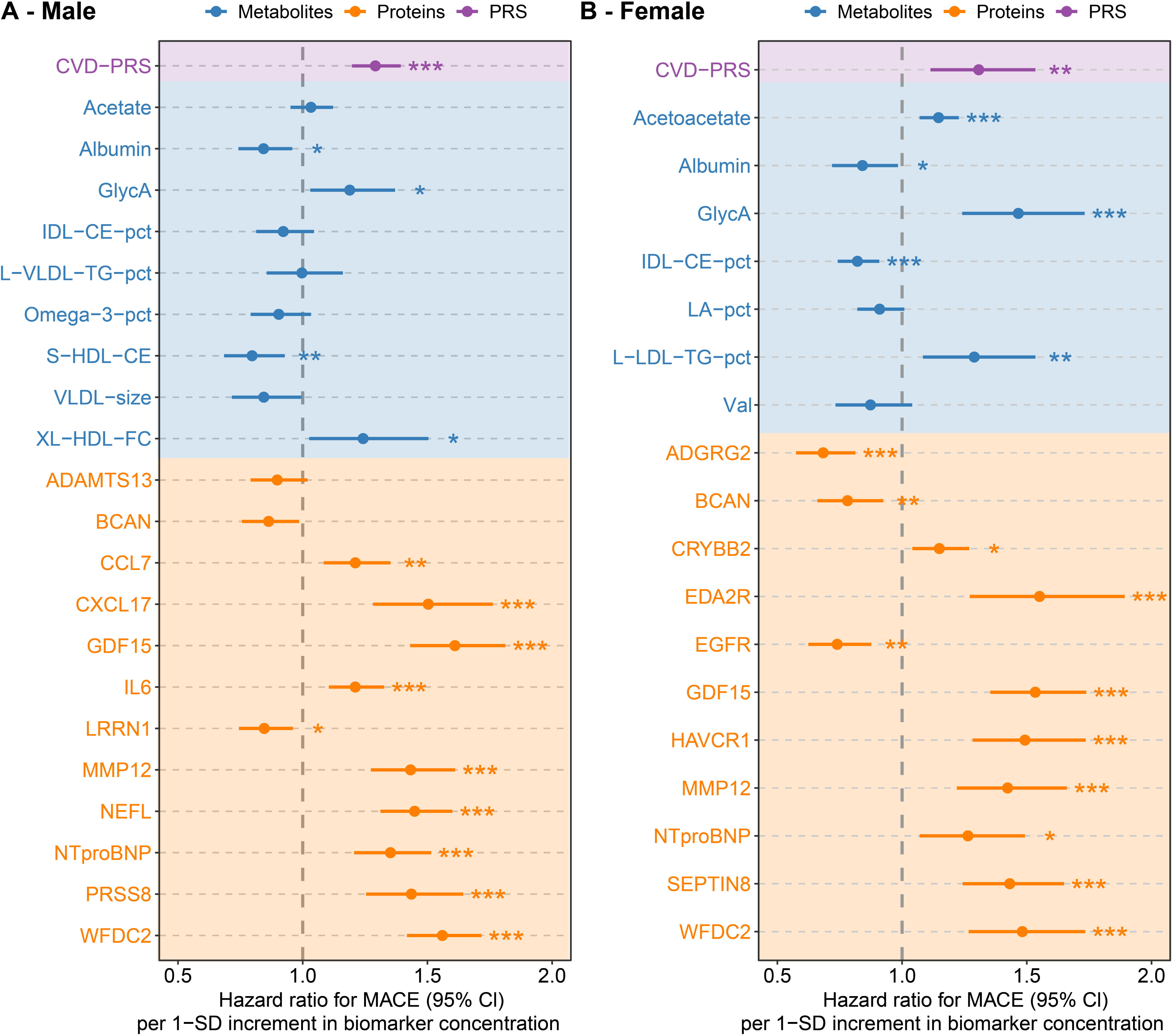
Associations of selected biomarkers with 10-year MACE risk in males and females in the validation set (N=7,214) Hazard ratios (HRs) and 95% confidence intervals for 1-SD increases in biomarker levels are shown for males (Panel A) and females (Panel B). **Abbreviations:** ADAMTS13, A disintegrin and metalloproteinase with thrombospondin motifs 13; BCAN, Brevican core protein; CCL7, C-C motif chemokine 7; CRYBB2, Beta-crystallin B2; CXCL17, C-X-C motif chemokine 17; EDA2R, Tumor necrosis factor receptor superfamily member 27; EGFR, Epidermal growth factor receptor; GDF15, Growth/differentiation factor 15; GlycA, Glycoprotein acetyls; IDL-CE-pct, The cholesteryl esters percentage of total lipids in intermediate-density lipoprotein; IL6, Interleukin-6; LA-pct, The linoleic acid percentage of total fatty acids; L-LDL-TG-pct, Triglycerides to total lipids in large low-density lipoprotein percentage; L-VLDL-TG-pct, The triglycerides percentage of total lipids in large very-low-density lipoprotein; LRRN1, Leucine-rich repeat neuronal protein 1; MMP12, Macrophage metalloelastase; NEFL, Neurofilament light polypeptide; NTproBNP, N-terminal prohormone of brain natriuretic peptide; Omega-3-pct, The Omega-3 fatty acids percentage of total fatty acids; PRS, polygenic risk score; PRSS8, Prostasin; S-HDL-CE, Cholesteryl esters in small high-density lipoprotein; SD, Standard deviation ; SEPTIN8, Septin-8; Val, Valine; VLDL-size, Average diameter for very-low-density lipoprotein particles; WFDC2, WAP four-disulfide core domain protein; XL-HDL-FC, Free cholesterol in very large high-density lipoprotein.

### MACE risk prediction by multi-omics

Figure 5 shows the predictive performance of the SCORE2 model extended with various multi-omics layers for 10-year MACE risk prediction. The addition of PRS, metabolomics, and proteomics each significantly improved model performance. The coefficients for all variables of the multi-omics extended models are detailed in **Supplemental Table S3**.

**Figure 5.**
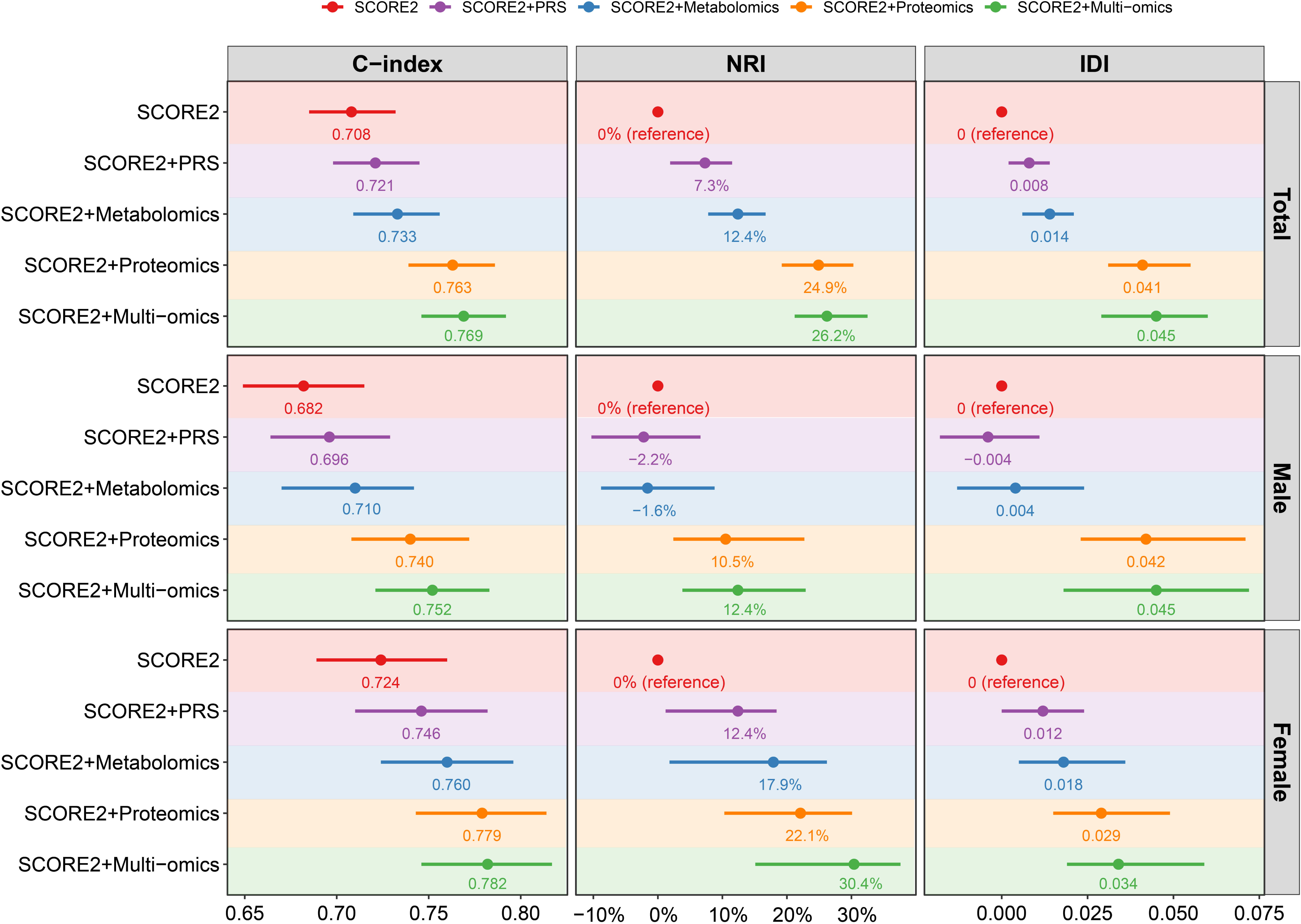
Performance of SCORE2 with and without omics data extensions for 10-year MACE prediction **Abbreviations**: IDI, Integrated discrimination index; NRI, Net reclassification index.

For the total population, integrating multi-omics data into the SCORE2 model significantly improved predictive performance, with the C-index increasing from 0.708 to 0.769 (ΔC-index = +0.061, *P*<0.001). The NRI was 26.2% (95% CI: 21.2%, 32.5%). In males, the C-index increased from 0.682 to 0.752 (ΔC-index = +0.070, *P*<0.001), with an NRI of 12.4% (95% CI: 3.8%, 22.9%). Similarly, in females, the C-index improved from 0.724 to 0.782 (ΔC-index = +0.058, *P*<0.001), with an NRI of 30.4% (95% CI: 15.1%, 37.6%). It was apparent that proteomics provided the most substantial improvement in both sexes, followed by metabolomics, and the PRS.

Figure 6 illustrates the incremental changes in C-index attributed to individual biomarkers in males (**6A**) and females (**6B**). Among all biomarkers, GDF15 contributed the most to risk prediction in both sexes, as reflected by the largest individual increase in the C-index when added to the SCORE2 model. Figures 6C and **6D** display the top 10 contributors to MACE risk prediction among males and females. For both sexes, the top 8 biomarkers were proteins, followed by the CVD-PRS and 1 metabolite, which underscores the high predictive ability of proteomic biomarkers.

**Figure 6.**
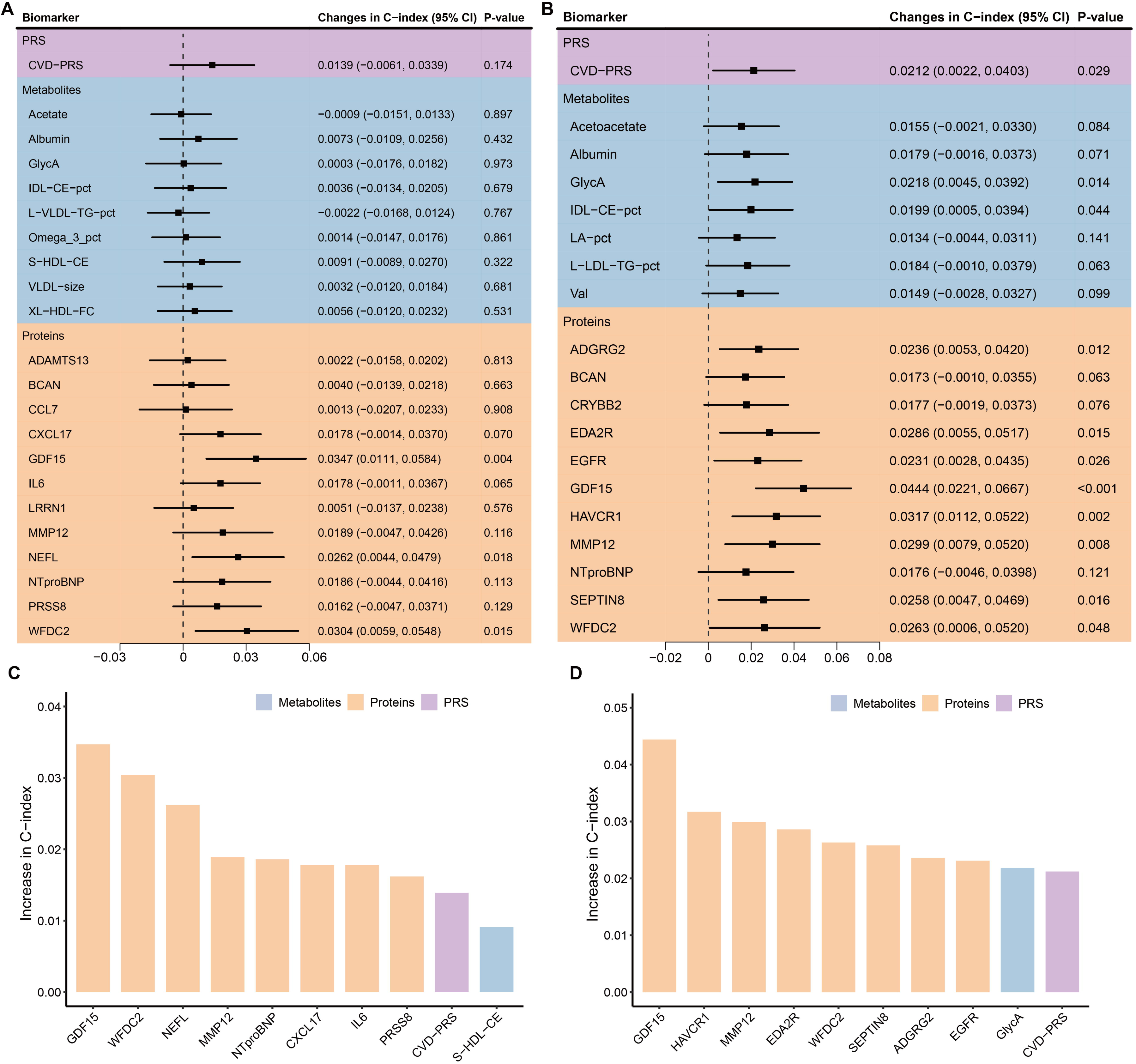
Incremental changes in C-index attributed to individual biomarkers Forest plots of the incremental change in C-index attributed to each biomarker for males (A) and females (B). Panels C and D show the top contributors in males and females, respectively. **Abbreviations:** ADAMTS13, A disintegrin and metalloproteinase with thrombospondin motifs 13; BCAN, Brevican core protein; CCL7, C-C motif chemokine 7; CRYBB2, Beta-crystallin B2; CXCL17, C-X-C motif chemokine 17; EDA2R, Tumor necrosis factor receptor superfamily member 27; EGFR, Epidermal growth factor receptor; GDF15, Growth/differentiation factor 15; GlycA, Glycoprotein acetyls; IDL-CE-pct, The cholesteryl esters percentage of total lipids in intermediate-density lipoprotein; IL6, Interleukin-6; LA-pct, The linoleic acid percentage of total fatty acids; L-LDL-TG-pct, Triglycerides to total lipids in large low-density lipoprotein percentage; L-VLDL-TG-pct, The triglycerides percentage of total lipids in large very-low-density lipoprotein; LRRN1, Leucine-rich repeat neuronal protein 1; MMP12, Macrophage metalloelastase; NEFL, Neurofilament light polypeptide; NTproBNP, N-terminal prohormone of brain natriuretic peptide; Omega-3-pct, The Omega-3 fatty acids percentage of total fatty acids; PRS, polygenic risk score; PRSS8, Prostasin; S-HDL-CE, Cholesteryl esters in small high-density lipoprotein; SD, standard deviation; SEPTIN8, Septin-8; Val, Valine; VLDL-size, Average diameter for very-low-density lipoprotein particles; WFDC2, WAP four-disulfide core domain protein; XL-HDL-FC, Free cholesterol in very large high-density lipoprotein.

In Figure 7, we evaluate which combination of omics data has the best model performance for MACE prediction. Among all single-omics extensions, proteomics provided the greatest improvement across the total population (C-index: 0.763 vs. 0.708; *P*<0.001), in males (0.740 vs. 0.682; *P*<0.001), and in females (0.779 vs. 0.724; *P*<0.001) and was therefore chosen as the next reference model. Adding the PRS to the proteomics extended SCORE2 model significantly further improved prediction among the total population (C-index: 0.769 vs. 0.763; *P*=0.007) and males (C-index: 0.749 vs. 0.740; *P*=0.017). The C-index also slightly improved among women but this difference was not statistically significant (C-index: 0.782 vs. 0.779; *P*=0.198). The addition of metabolomics yielded no further benefit. Thus, the best model performance was observed for a combination of the variables of the SCORE2 model, proteomics data, and the CVD-PRS. **Supplemental Table S4** presents the sex-specific ß-coefficients for this optimal multi-omics combination model. Comparing this model with a metabolomics extended version confirmed that metabolomics did not further improve its predictive abilities (Figure 7).

**Figure 7.**
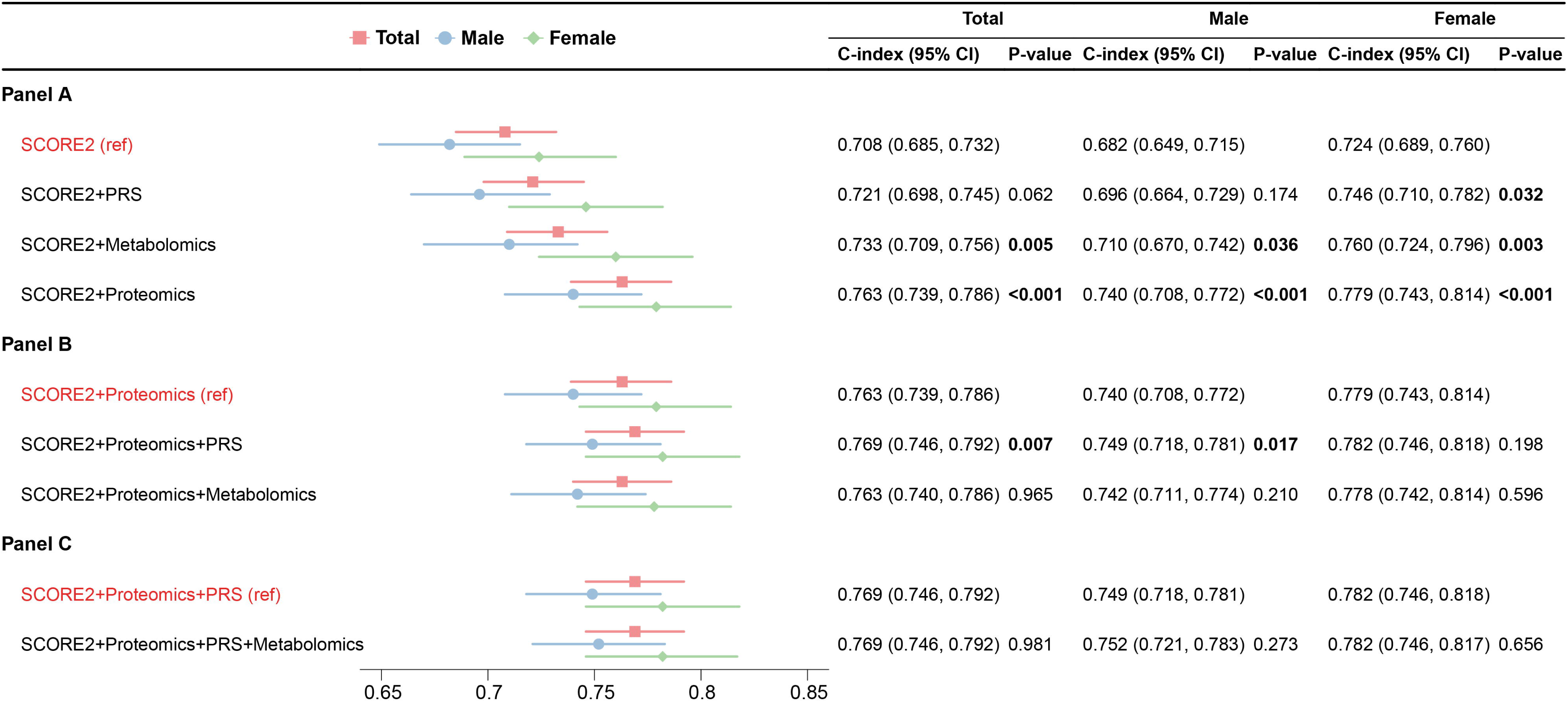
C-index comparison of single-omics and multi-omics extensions to SCORE2 in MACE prediction Panel A shows the C-index with 95% confidence intervals for models extending SCORE2 with individual omics layers (PRS, metabolomics, or proteomics). Panel B compares multi-omics combinations using SCORE2+proteomics as the reference.

**Supplemental Figure S1**. presents the risk reclassification tables for low-risk (≤7.5%), intermediate-risk (>7.5-15%), and high-risk (>15%) categories. Adding all multi-omics data to the SCORE2 model significantly improved the NRI in males (12.4%) and females (30.4%). The optimal multi-omics combination model, which excludes metabolomics, achieved comparable NRIs of 12.0% in males and 28.6% in females. **Supplemental Figure S2** displays the calibration curves for the SCORE2 model and its multi-omics extensions. The optimal multi-omics combination model demonstrated good model calibration.

## Discussion

This study investigated the integration of multi-omics data into the SCORE2 model to improve 10-year risk prediction of MACE. By applying sex-specific biomarker selection, we developed novel risk algorithms for women and men, incorporating a CVD-PRS, metabolites, and proteins, which yielded significant improvements in predictive performance. The optimal model was the combination of proteomics and the CVD-PRS with the SCORE2 model, which achieved predictive accuracy comparable to that of the full multi-omics model in both males and females. These findings highlight the importance of integrating multi-omics data and sex-specific modeling in enhancing precision cardiovascular risk prediction.

To our knowledge, this is the first study to investigate the predictive value of multi-omics data (PRS, metabolomics, and proteomics) integrated into an established cardiovascular risk model.

Previous studies have reported improvements in cardiovascular risk prediction using single omics layers [9, 10, 14, 32–37]. For example, Li et al. demonstrated that combining a CVD-PRS with the SCORE2 model significantly enhanced risk stratification in the UK Biobank population [32]. A systematic review and meta-analysis analyzing 21 studies with 54,337 participants reported an average increase of 0.04 in the C-index when incorporating metabolomics into cardiovascular risk models [35]. Similarly, in an Icelandic cohort, an increase in the C-index by 0.014 was observed for predicting MACE when a protein risk score was added to a traditional risk model [36].

Our study showed for the first time that integrating both proteomics and CVD-PRS data is useful to enhance cardiovascular risk prediction by a traditional risk factor model, while metabolomics data add too little C-index improvement to be of clinical relevance after proteomics data were added. Among the individual omics layers, proteomics contributed the greatest predictive value, outperforming both the CVD-PRS and metabolomics in both males and females. Even the addition of the CVD-PRS to a proteomics extended SCORE2 model added little incremental predictive value (although it was statistically significant), which likely makes the proteomics extended SCORE2 model the model with the highest translation potential because it is almost as good as the model additionally including the CVD-PRS and does not require cost-intensive genetic testing and the labor of using software to compute the CVD-PRS from several million SNPs (Single nucleotide polymorphisms). While several studies have tested large-scale omics features comprising hundreds of proteins or metabolites, the costs and logistical demands of validating and harmonizing such extensive assays limit their feasibility for clinical translation [38, 39]. Our protein-panel only includes 12 proteins for men and 11 proteins for women, which could be measured at lower costs than full proteomics. However, to measure these few proteins simultaneously with absolute concentrations, the assays need to be developed first with, for example, the OLINK Focus technology, which can measure up to 21 proteins from the whole OLINK Explore protein library (https://olink.com/products/olink-focus). Subsequently, the predictive performance of this OLINK Focus panel for 10-year MACE risk should be validated in external cohorts before this sex-specific prediction algorithm could be used in the clinical routine.

Sex differences in cardiovascular risk and multi-omics biomarkers have been well-documented [16, 17, 40, 41]. Previous studies typically included sex as a covariate in the model derivation process rather than selecting biomarkers separately for males and females. We addressed these limitations by developing sex-specific sparse models, restricting the tested omics features to a small number of highly informative biomarkers. Our previous work has validated the effectiveness of this approach in different populations [13, 14]. For instance, Ho et al. reported that incorporating 85 proteins into the PREVENT risk model improved the C-index for predicting MACE by 0.054 (from 0.745 to 0.799) and achieved a categorical NRI of 9% [37]. Similarly, Royer et al. found that adding 114 proteins to the SCORE2 model increased the AUC by 0.031 (from 0.740 to 0.771) and resulted in an NRI of 14.0% [10]. Compared to these studies, our sex-specific sparse model achieved a greater improvement in model discrimination (C-index increased from 0.716 to 0.778) [13]. Notably, this enhancement was achieved by incorporating only 12 proteins for males and 11 for females into the SCORE2 model, rather than the 85 proteins used by Ho et al. or the 114 proteins used by Royer et al. Furthermore, the NRI observed in our analysis (19.9%) was higher than that reported by Ho et al. (9%) and Royer et al. (14.0%), highlighting the effectiveness of a sex-specific approach in improving predictive accuracy. By integrating sex-specific biomarker selection while accounting for baseline differences in cardiovascular risk profiles, our model facilitates more personalized and clinically actionable risk stratification. In addition, measuring up to 12 proteins is more cost-effective and practical for clinical implementation compared to measuring 114 proteins, facilitating the translation of our approach into routine clinical practice.

This study’s strengths include the availability of multi-omics data from all 3 layers (genetics, proteomics, and metabolomics) in a large cohort study with 10-year MACE follow-up. The use of sex-specific modelling considered the biological differences between males and females and is a first step towards personalized CVD prevention. However, several limitations should be noted. First, the metabolomics platform of Nightingale Health is limited to 250 metabolites and more comprehensive, untargeted metabolomics data measured with mass spectrometry methods might have yielded better results. Second, our analyses were primarily based on middle-aged and older adults of European ancestry, necessitating validation in more diverse populations with varying baseline cardiovascular risks and ethnic backgrounds, to ensure the generalizability of our findings. Third, although we provided beta coefficients for selected biomarkers, these coefficients may require recalibration when applied to populations with varying demographic and clinical profiles or when using different measurement platforms. Finally, the study did not include an in-depth cost-effectiveness analysis of integrating multi-omics data into clinical practice, which is crucial for assessing the feasibility of large-scale implementation.

## Conclusion

This study demonstrates for the first time in the same data set that adding a CVD-PRS, metabolomics, and proteomics data individually to the SCORE2 model significantly improves its ability for 10-year MACE prediction, with the proteomics data clearly outperforming the other omics data. The optimal omics data combination was proteomics and the CVD-PRS, while metabolomics did not improve the model further. Our findings underscore the added predictive value of multi-omics data in refining cardiovascular risk assessment. However, for better translation into clinical routine, our results also show that it would be possible to measure just a small set of proteins specifically tailored to men and women (up to 12) and obtain an almost as good model discrimination as a model with full multi-omics data.

## Non-standard Abbreviations and Acronyms

CI: Confidence intervals
CVD: Cardiovascular disease
CXCL17: C-X-C motif chemokine 17
GDF15: Growth/differentiation factor 15
GlycA: Glycoprotein acetyls
GWAS: Genome-wide association studies
HDL-C: High-density lipoprotein cholesterol
HR: Hazard ratio
IDI: Integrated discrimination index
IDL-CE-pct: The cholesteryl esters percentage of total lipids in intermediate-density lipoprotein
IL6: Interleukin-6
LA-pct: The linoleic acid percentage of total fatty acids
LASSO: Least absolute shrinkage and selection operator
L-LDL-TG-pct: Triglycerides to total lipids in large low-density lipoprotein percentage
MACE: Major adverse cardiovascular events
NMR: Nuclear magnetic resonance
NRI: Net reclassification index
PRS: Polygenic risk scores
SBP: Systolic blood pressure
SD: Standard deviation
SNPs: Single nucleotide polymorphisms
UKB: UK Biobank
Val: Valine
WFDC2: WAP four-disulfide core domain protein 2

## Declarations

### Author contributions

R.X. and B.S. generated the idea for the study and formulated the analytical plan. R.X. performed the data analyses and drafted the manuscript. B.S. revised it. M.B., S.S., L.P., T.V., and H.B. contributed to the interpretation of the data and provided valuable intellectual content to the discussion. All authors critically reviewed the manuscript and approved the final version. The corresponding author attests that all listed authors meet authorship criteria and that no others meeting the criteria have been omitted. R.X. and B.S. had full access to UK Biobank data used for this study and are the guarantors of the manuscript and accept full responsibility for the work and/or the conduct of the study.

## Acknowledgements

This research has been conducted using the UK Biobank Resource under Application Number 101633. This work uses data provided by patients and collected by the NHS as part of their care and support. We would like to thank all participants of the UK Biobank as well as the staff of the UK Biobank assessment centers for their contributions.

## Human Ethics and Consent to Participate declarations

The study was conducted in accordance with the Declaration of Helsinki. The UK Biobank study has obtained approval from the North West Multi-centre Research Ethics Committee (MREC) (reference 11/NW/0382). All participants provided written informed consent.

## Data availability

The data that support the findings of this study are available from UK Biobank (https://www.ukbiobank.ac.uk/) but restrictions apply to the availability of these data, which were used under license for the current study (Application Number 101633), and so are not publicly available. Data are however available from the authors upon reasonable request and with permission of UK Biobank.

