## Supplemental materials for "Sex-specific prediction of major cardiovascular events in apparently healthy individuals with multi-omics data"

**Supplemental Table S1.** Selected multi-omic biomarkers

| **Biomarkers** | **Males** | **Females** | **Reference** |
| --- | --- | --- | --- |
| PRS | Standard PRS for CVD | Standard PRS for CVD | The standard CVD-PRS was derived using a genome-wide meta-analysis of 9 external GWAS datasets comprising 1,840,289 individuals and has been previously published [1]. |
| Metabolites | Acetate, Albumin, GlycA, IDL-CE-pct, L-VLDL-TG-pct, Omega-3-pct, S-HDL-CE, VLDL-size, XL-HDL-FC | Acetoacetate, Albumin, GlycA, IDL-CE-pct, LA-pct, L-LDL-TG-pct, Val | The metabolites were selected using sex-specific LASSO regression in the UKB (n=130,927). See previous work [2]. |
| Proteins | ADAMTS13, BCAN, CCL7, CXCL17, GDF15, IL6, LRRN1, MMP12, NEFL, NTproBNP, PRSS8, WFDC2 | BCAN, CRYBB2, EDA2R, EGFR, GDF15, HAVCR1, MMP12, NTproBNP, SEPTIN8, WFDC2 | The proteins were selected using sex-specific LASSO regression in the UKB (n=33,166). See previous work [3]. |

**Abbreviations:** ADAMTS13, A disintegrin and metalloproteinase with thrombospondin motifs 13; BCAN, Brevican core protein; CCL7, C-C motif chemokine 7; CRYBB2, Beta-crystallin B2; CXCL17, C-X-C motif chemokine 17; EDA2R, Tumor necrosis factor receptor superfamily member 27; EGFR, Epidermal growth factor receptor; GDF15, Growth/differentiation factor 15; GlycA, Glycoprotein acetyls; IDL-CE-pct, The cholesteryl esters percentage of total lipids in intermediate-density lipoprotein; IL6, Interleukin-6; LA-pct, The linoleic acid percentage of total fatty acids; L-LDL-TG-pct, Triglycerides to total lipids in large low-density lipoprotein percentage; L-VLDL-TG-pct, The triglycerides percentage of total lipids in large very-low-density lipoprotein; LRRN1, Leucine-rich repeat neuronal protein 1; MMP12, Macrophage metalloelastase; NEFL, Neurofilament light polypeptide; NTproBNP, N-terminal prohormone of brain natriuretic peptide; Omega-3-pct, The Omega-3 fatty acids percentage of total fatty acids; PRS, polygenic risk score; PRSS8, Prostasin; S-HDL-CE, Cholesteryl esters in small high-density lipoprotein; SEPTIN8, Septin-8; Val, Valine; VLDL-size, Average diameter for very-low-density lipoprotein particles; WFDC2, WAP four-disulfide core domain protein; XL-HDL-FC, Free cholesterol in very large high-density lipoprotein.

**Supplemental Table S2.** Definition of endpoint major cardiovascular event (MACE)

| **Fatal MACE – cause-specific mortality due to any of the following:** | |
| --- | --- |
| *Endpoints included* | *ICD10-codes* |
| Hypertensive disease | I10-16 |
| Ischemic heart disease | I20-25 |
| Arrhythmias, heart failure | I46-52 |
| Cerebrovascular disease | I60-69 |
| Atherosclerosis/aortic aneurysm | I70-73 |
| Sudden death and death within 24 hours of symptom onset | R96.0-96.1 |
| *Endpoints excluded from the above endpoint:* | *ICD10-codes* |
| Myocarditis, unspecified | I51.4 |
| Subarachnoid haemorrhage | I60 |
| Subdural hemorrhage | I62 |
| Cerebral aneurysm | I67.1 |
| Cerebral arteritis | I68.2 |
| Moyamoya | I67.5 |
| **Non-fatal MACE** | *ICD10-codes* |
| Non-fatal myocardial infarction | I21-I23 |
| Non-fatal stroke | I61, I63-I66, I69 |

### **Supplemental** **Table S3.** ß-coefficients of the variables of the SCORE2 model extended by multi-omics for 10-year prediction of major cardiovascular events

| **Risk factor (units)** | **Coefficient** | |
| --- | --- | --- |
|  | **Male** | **Female** |
| **SCORE2 variables** |  |  |
| Age (per 5 years) | 0.1248 | 0.3475 |
| Current smoking | -0.0638 | 0.5276 |
| Systolic blood pressure (per 20mmHg) | 0.0710 | 0.2138 |
| Total cholesterol (per 1 mmol/L) | 0.0728 | 0.0370 |
| HDL cholesterol (per 0.5 mmol/L) | -0.0004 | -0.1207 |
| Smoking interaction with age | -0.0223 | -0.1793 |
| SBP interaction with age | -0.1158 | -0.0273 |
| Total cholesterol interaction with age | -0.0116 | -0.0449 |
| HDL interaction with age | 0.0099 | -0.0325 |
| **Additional PRS** **(per 1 SD)** |  |  |
| CVD-PRS | 0.2114 | 0.1476 |
| **Additional metabolites** **(per 1 SD)** |  |  |
| Acetate | -0.0247 | - |
| Acetoacetate | - | 0.0017 |
| Albumin | -0.0093 | -0.0656 |
| GlycA | 0.0304 | 0.0180 |
| IDL-CE-pct | -0.0130 | -0.0916 |
| LA-pct | - | 0.0041 |
| L-LDL-TG-pct | - | 0.0061 |
| L-VLDL-TG-pct | -0.0522 | - |
| Omega-3-pct | -0.0466 | - |
| S-HDL-CE | -0.0436 | - |
| Val | - | -0.0276 |
| VLDL-size | -0.0387 | - |
| XL-HDL-FC | 0.0640 | - |
| **Additional proteins** **(per 1 SD)** |  |  |
| ADAMTS13 | -0.1388 | - |
| ADGRG2 | - | -0.1080 |
| BCAN | -0.1774 | -0.0816 |
| CCL7 | 0.1626 | - |
| CRYBB2 | - | 0.0921 |
| CXCL17 | 0.0742 | - |
| EDA2R | - | 0.0325 |
| EGFR | - | -0.0822 |
| GDF15 | 0.1704 | 0.1990 |
| HAVCR1 | - | 0.0648 |
| IL6 | -0.0039 | - |
| LRRN1 | -0.0892 | - |
| MMP12 | 0.2227 | 0.0713 |
| NEFL | 0.1276 | - |
| NTproBNP | 0.2052 | 0.1898 |
| PRSS8 | 0.1012 | - |
| SEPTIN8 | - | 0.1523 |
| WFDC2 | 0.0435 | 0.2055 |

**Abbreviations:** ADAMTS13, A disintegrin and metalloproteinase with thrombospondin motifs 13; BCAN, Brevican core protein; CCL7, C-C motif chemokine 7; CRYBB2, Beta-crystallin B2; CXCL17, C-X-C motif chemokine 17; EDA2R, Tumor necrosis factor receptor superfamily member 27; EGFR, Epidermal growth factor receptor; GDF15, Growth/differentiation factor 15; GlycA, Glycoprotein acetyls; IDL-CE-pct, The cholesteryl esters percentage of total lipids in intermediate-density lipoprotein; IL6, Interleukin-6; LA-pct, The linoleic acid percentage of total fatty acids; L-LDL-TG-pct, Triglycerides to total lipids in large low-density lipoprotein percentage; L-VLDL-TG-pct, The triglycerides percentage of total lipids in large very-low-density lipoprotein; LRRN1, Leucine-rich repeat neuronal protein 1; MMP12, Macrophage metalloelastase; NEFL, Neurofilament light polypeptide; NTproBNP, N-terminal prohormone of brain natriuretic peptide; Omega-3-pct, The Omega-3 fatty acids percentage of total fatty acids; PRS, polygenic risk score; PRSS8, Prostasin; S-HDL-CE, Cholesteryl esters in small high-density lipoprotein; SD, standard deviation; SEPTIN8, Septin-8; Val, Valine; VLDL-size, Average diameter for very-low-density lipoprotein particles; WFDC2, WAP four-disulfide core domain protein; XL-HDL-FC, Free cholesterol in very large high-density lipoprotein.

**Supplemental Table S4.** ß-coefficients of the variables of the optimal multi-omics combination models for 10-year prediction of major cardiovascular events

| **Risk factor (units)** | **ß coefficients** | |
| --- | --- | --- |
|  | **Male** | **Female** |
| **SCORE2 variables** |  |  |
| Age (per 5 years) | 0.1115 | 0.3393 |
| Current smoking | -0.0499 | 0.5188 |
| Systolic blood pressure (per 20mmHg) | 0.0677 | 0.2107 |
| Total cholesterol (per 1 mmol/L) | 0.0773 | 0.0119 |
| HDL cholesterol (per 0.5 mmol/L) | 0.0008 | -0.1737 |
| Smoking interaction with age | -0.0212 | -0.1711 |
| SBP interaction with age | -0.0697 | -0.0255 |
| Total cholesterol interaction with age | -0.0133 | -0.0447 |
| HDL interaction with age | 0.0186 | -0.0330 |
| **Additional PRS** **(per 1 SD)** |  |  |
| CVD-PRS | 0.2107 | 0.1478 |
| **Additional proteins** **(per 1 SD)** |  |  |
| ADAMTS13 | -0.1468 | - |
| ADGRG2 | - | -0.1169 |
| BCAN | -0.1749 | -0.0881 |
| CCL7 | 0.1733 | - |
| CRYBB2 | - | 0.0901 |
| CXCL17 | 0.0745 | - |
| EDA2R | - | 0.0345 |
| EGFR | - | -0.0941 |
| GDF15 | 0.1903 | 0.2186 |
| HAVCR1 | - | 0.0808 |
| IL6 | 0.0090 | - |
| LRRN1 | -0.0945 | - |
| MMP12 | 0.2263 | 0.0720 |
| NEFL | 0.1278 | - |
| NTproBNP | 0.2139 | 0.1931 |
| PRSS8 | 0.0639 | - |
| SEPTIN8 | - | 0.1662 |
| WFDC2 | 0.0671 | 0.2129 |

**Abbreviations:** ADAMTS13, A disintegrin and metalloproteinase with thrombospondin motifs 13; BCAN, Brevican core protein; CCL7, C-C motif chemokine 7; CRYBB2, Beta-crystallin B2; CXCL17, C-X-C motif chemokine 17; EDA2R, Tumor necrosis factor receptor superfamily member 27; EGFR, Epidermal growth factor receptor; GDF15, Growth/differentiation factor 15; IL6, Interleukin-6; LRRN1, Leucine-rich repeat neuronal protein 1; MMP12, Macrophage metalloelastase; NEFL, Neurofilament light polypeptide; NTproBNP, N-terminal prohormone of brain natriuretic peptide; PRS, polygenic risk score; PRSS8, Prostasin; SD, standard deviation; SEPTIN8, Septin-8; WFDC2, WAP four-disulfide core domain protein.

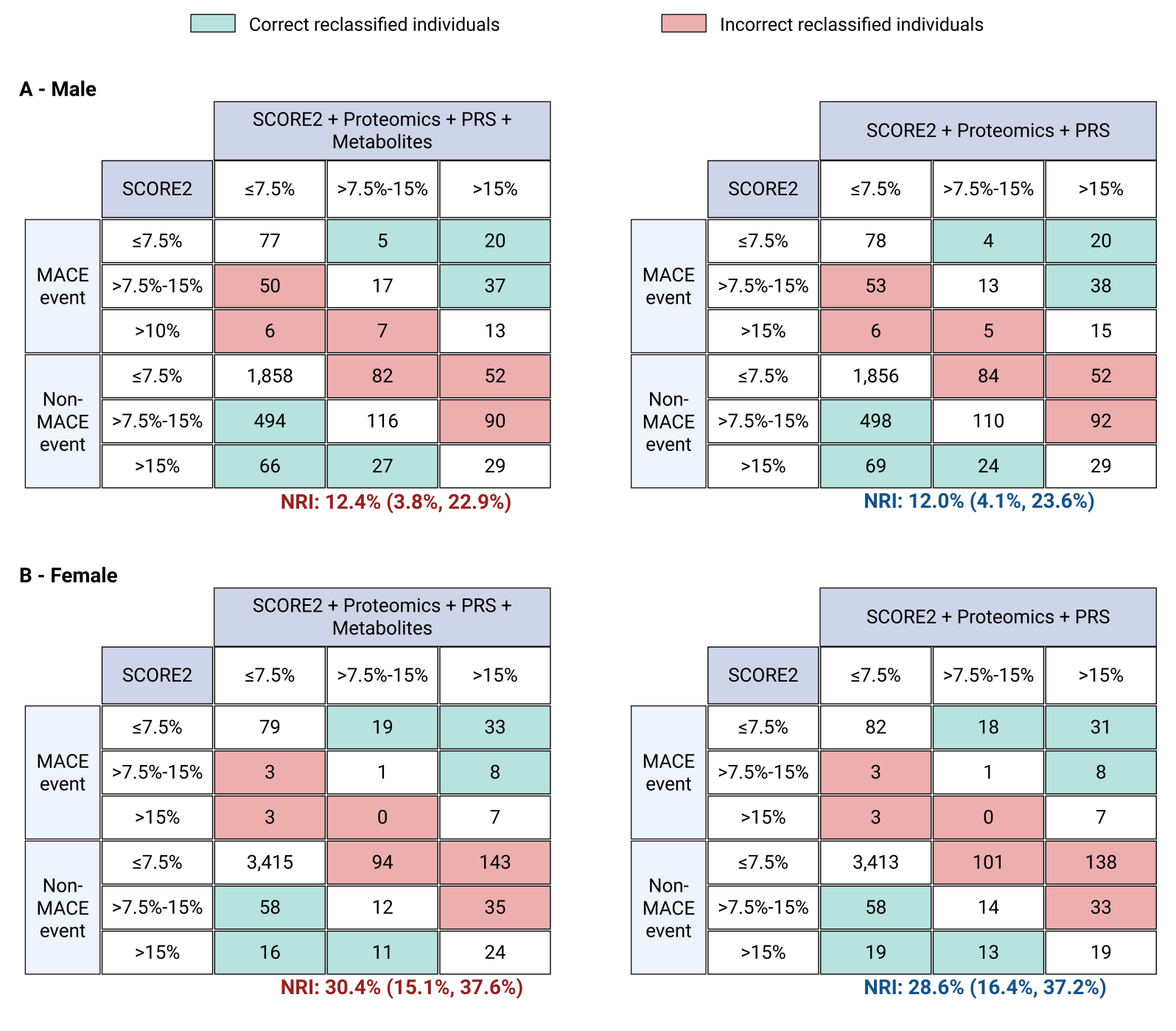

### **Supplemental Figure S1.** Reclassification table of the SCORE2 model with Proteomics + PRS + Metabolomics or optimal multi-omics combination in the validation set (30% of UK Biobank, N=7,214)

The reclassification of individuals into cardiovascular risk categories (≤7.5%, >7.5%-15%, >15%) for major adverse cardiovascular events (MACE) using the SCORE2 model alone and with the addition of proteomic biomarkers. The results are shown for the males (**Panel A**) and females (**Panel B**).

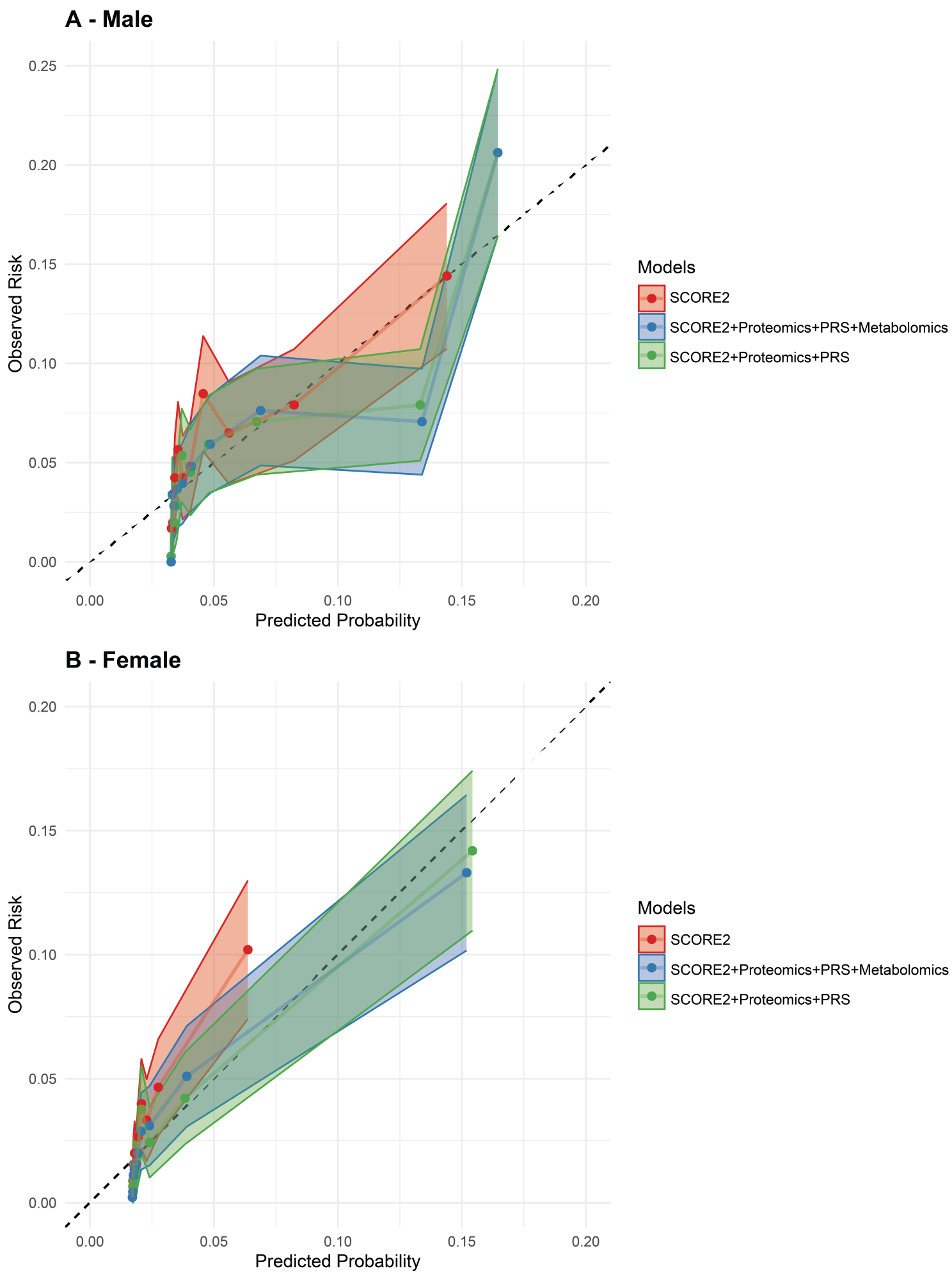

**Supplemental Figure S2.** Calibration curves of the SCORE2 model with and without Proteomics + PRS + Metabolomics or optimal multi-omics combination extended for 10-year MACE risk prediction in the validation set (30% of UK Biobank, N=7,214)
